# Persistence of venom-induced consumption coagulopathy (VICC) associated with the late application of antivenom after snakebite by Bothrops complex: a retrospective cohort study

**DOI:** 10.64898/2026.01.02.26343340

**Authors:** Ana M. Parada, María A. Arteaga, Lina M. Peña-Acevedo, Víctor De La Espriella, Diego E. Giraldo, Johan S. Morales, Gilma N. Hernandez, Carlos E. Vallejo-Bocanumen

## Abstract

**Introduction:** Antivenom neutralizes the effects of snake venom toxins. Its timely administration can by critical in reducing the risk of complications, including serious clinical hemorrhage. The objective of this study was to evaluate the relationship between the late administration of antivenom and the persistence of venom-induced consumption coagulopathy (VICC) in patients with Bothrops snakebite envenomation.

**Methods:** We conducted a retrospective observational study in three hospitals referent in the management of snakebite envenomation in a region of Colombia. Patients were identified through ICD10 codes. The information was extracted from patients’ clinical records. Late and early antivenom administration patients were compared; “late” was defined as a period between snakebite and antivenom administration >4h. Definition of persistence of VICC was as an International Normalized Ratio >1.5 between 25-72h after snakebite. The clinical bleeding (as reported by clinicians) was also explored. Groups were compared through *X^2^* of association; binary and multiple logistic regressions were ran to estimate the magnitude of the crude and adjusted relative risk (RR) for the analyzed outcomes.

**Results:** 304 patients with snakebite from *Bothrops spp.*were analyzed; 138 (45.3%) with late antivenom administration, and 166 (54.7%) early. The persistence of VICC was more frequent in the late antivenom group (2.06 [95% IC 1.11-3.83]), as well as for clinical bleeding and major bleeding (1.97[1.30-2.98]; 3.00 [1.19-7.54], respectively).

**Discussion:** Late antivenom administration was associated with increased risk in the persistence of VICC and major bleeding in patients with envenomation by a Bothrops complex snakebite. The results reported in the literature are diverse; inter and intra species variability and definitions can influence the finding.

**Conclusions:** The Timely administration of antivenom was associated with an increased risk in persistence of VICC.

**Synopsis:** Snakebites can cause different levels of damage to humans, depending on the type of venom. The venom produced by pit vipers, specifically those of the genus Bothrops, can change blood clotting and cause bleeding that can be life-threatening. Most snakebites from these vipers occur in tropical areas where the most affected are farmers and rural people. There is often a lack of access to antivenom treatment, depending on access to and provision of health services. It seems that giving the antivenom at the right time can reduce the risk of complications. However, it is still unclear whether waiting to give the antivenom increases the risk of continued envenoming and bleeding. This study shows that waiting to treat a patient can lead to a higher chance of bleeding. This information can help doctors, decision makers, and lawmakers improve treatment options and access in remote areas.

## Introduction

Snakebite is classified as a neglected tropical disease by the WHO. Annually, 4.5–5.4 million individuals worldwide experience a snakebite [1], [2] of which 38,900 – 78,600 die resulting in 38 years of life lost due to disability per 100,000 inhabitants[3]. In Colombia, nearly 5,000 snakebite cases are reported per year, with an incidence rate of 9.4–11 cases per 100,000 inhabitants. Of these cases, 68% were attributed to pit vipers. The reported mortality rate was 0.6%[4].

Venom-induced consumption coagulopathy (VICC) is a prevalent and important hemotoxicity in pit viper envenomation syndrome, it manifests as coagulopathy and can progress to life-threatening bleeding. Antivenom administration is currently the treatment of choice, which neutralizes the circulating toxins and thus facilitates the recovery of VICC [5], [6], [7].

The hemotoxicity has been attributed characteristics of the species involved in the snakebite. However, factors inherent to the socio-cultural context can also play a role [2], [7], [8]. Some studies suggest that antivenom administration before two hours or six hours after the snakebite could be associated with an improved recovery of VICC, however there also have been described a persistence of VICC in late administration of the antivenom [6], [9], [10], [11], which suggests a time-dependent phenomenon, but the role that time plays in its administration remains ambiguous.

In Colombia, pit viper bites, particularly of the Bothrops species, account for more than two-thirds of all reported snakebites, and these are associated with high morbidity, and therefore, a public health concern [1], [4]. Snakebite envenomation management is based on the Toxicological Emergencies Guide of the Colombian Ministry of Health, that guides medical interventions with range of antivenoms available for its treatment [12]. However, 70% of the events occur in rural areas with geographical barriers that limit the adequate provision of health services, thereby making access to a comprehensive care, including antivenoms, more difficult, which could increase the timespan to the antivenom [13].

The aim of this study was to evaluate the impact of the early antivenom administration on the persistence of VICC in the population with Bothrops snakebite envenomation.

## Methods

### Study Design, Setting and Participants

We carry out a cohort study in two high-complexity hospitals in the city of Medellín, Colombia. They served as referral institutions in the northwestern region of the country for the management of patients with snakebite envenomation. They had specialist (including clinical toxicologists), protocols and services (clinical laboratory, blood bank) for the comprehensive management of this condition.

The patients included in this study were those admitted to the referral hospitals between January 1, 2017 and December 31, 2022 with Bothrops complex snakebite envenomation [14], who had received at least one dose of the antivenom after a positive 20 minutes whole blood clothing time (20WBCT) recorded in the medical records of the hospital from which they were transferred.

Patients with clinical records without information to determine the time between the snakebite and the antivenom application, elapid snakebites cases, and patients with a history of coagulation disorders or being treated with anticoagulants at the time of the snakebite were excluded.

### Data collection and classification

Patients were identified through review of the institutional database using the ICD-10 diagnostic code (T630: *Toxic effect of contact with poisonous animals: snake venom*). All medical records with this code were reviewed, and inclusion and exclusion criteria were then determinate.

The data were obtained from the digital medical records of both hospitals, and then, entered into a form designed for this study based on a Google forms hosted in the cloud. The information was collected in a Microsoft Excel^®^ (version 14.1.0, Redmond, Washington, USA: Microsoft, 2011) spreadsheet for storage and subsequent processing and analysis. A pilot test was conducted with 10 clinical histories, which allowed refinement of the collection instrument.

Data of the following variables were collected: age; gender, health care affiliation regime, region of origin (municipality and department), the characteristics of the snakebite including registered timings of: snakebite; medical assistance; and antivenoms administration. The coagulation tests and other laboratory results (ionogram, liver and kidney function, CK, blood count, acute phase reactants), the insufficient antivenom dosage according to the severity classification of the viperid envenomation, and the type of prehospital nonmedical management received, were also documentated.

Clinical observation and review of coagulation test and other laboratory results were collected in the first three days since the admission. In all cases, first coagulation test was taken at high-complexity hospital admission, and subsequent measurements were done at discretion of the medical staff.

Snakebite envenoming severity was classified according to Toxicological Emergencies Guide of the Colombian Ministry of Health, based on information of snake identification according to the specimen itself or the information provided by the patient or proxies, and the presence of local and systemic findings, including local (pain, edema, blisters and local bleeding) and systemic (VICC, shock and rhabdomyolysis) findings. The presence of local findings in more than three body segments or the trunk, or systemic involvement such as hemorrhage in the nervous system or VICC, renal failure, or shock, was defined as severe envenoming; bleeding without systemic involvement or VICC, and local findings in two body segments was considered moderate envenoming; the remainder was considered mild envenoming.[12].

Antivenom brands available in Colombia for snakebite envenoming treatment: Suero Antiofídico Polivalente^®^, Suero Antiofídico Polivalente Liofilizado^®^ both brands produced domestically by Instituto Nacional de Salud de Colombia (public), Probiol^®^ and Antivipmyn TRI^®^ by Silanes) were also registered. The antivenom dose was calculated by the severity of the poisoning, the specific brand and the neutralizing capacity of each antivenom [12].

Late antivenom administration was defined as a period between snakebite and antivenom administration >4 hours, and ≤4 hours as early. This threshold was established based on previous studies that defined antivenom administration less than 2 or 6 hours after snakebite as early [9], [10], [15]. For our study, the research team defined an intermediate of four hours. The timespan between the snakebite and the antivenom administration was defined as the difference between the two, in minutes, according to the clinical record.

Clinical bleeding was defined as any kind of bleeding (through skin, epistaxis, gingivorrhagia, gastrointestinal loss, urine samples, punctures or wounds; or in central nervous system documented by a head CT) observed by health staff and recorded in medical history; major bleeding was defined as Intracerebral hemorrhage or a documented level of hemoglobin lesser than 8 grams per deciliter with or without an observed hemorrhage reported in the first 72 hour after hospital admission; rhabdomyolysis were defined as a CK level > 1000; acute kidney injury was defined by KDIGO criteria based on the laboratory data and information provided in the clinical record.

VICC was defined as an INR >1.5 at measured at any moment during the first 72 hours after hospital admission [16].

### Outcomes

The main outcome was persistence of VICC, defined as a high INR >1.5 measured between 25 and 72 hours post-bite [16]. We consider this time period reasonable for VICC, as patients in this time period could persist with abnormal coagulation tests, as reported in the literature. [16].

### Sample size and sampling

The sample size was calculated from a reference Group from a study conducted in a hospital population [13], and from studies [15] [10] that evaluated the relationship between the time of application and the development of complications. With a power of 80%, a confidence level of 95%, a risk of 28.4% in the unexposed group and 31.4% in the exposed group and a ratio 1:1 between exposed and non-exposed patients, a sample of 290 patients was estimated, 145 patients in each group [17].

A non-probabilistic, convenience and sequential sampling was carried. Patients were followed from admission to hospital discharge.

### Ethical aspects

This research was classified as *no risk* by Colombian regulatory laws (Resolution Number 8430 of 1993 from the Colombian Ministry of Health). The approval of the project and the waiver of informed consent to participate in the study were granted by the Research Ethics Committee of the institutions participating in the study, through Minutes Number 06-2022 dated February 25, 2022, and March 5, 2022. This study was conducted in accordance with tenets of the Helsinki Declaration.

### Statistical analysis

Descriptive statistics were used for the socio-demographic variables and clinical characteristics of the patients. Absolute and relative frequencies were calculated for the qualitative variables. For quantitative variables, the mean and standard deviation (SD), or the median and interquartile range (IQR) were reported, depending on distribution of the data. Multiple histograms and the Kolmogorov-Smirnov test were used to determine the distribution of data in the late and early groups.

VICC frequency was estimated as the proportion of patients with an INR >1.5 over the population in each group, in the first 24 hours and between 25 and 72 hours post-bite.

To determine the differences in the proportions of primary and secondary outcomes between both groups, the *X^2^* test was used. To estimate the magnitude of the association between late antivenom administration and the persistence of VICC, binary logistic regression models were performed; multivariate logistic regression were performed to adjust the models with potential confounders.

The insufficient antivenom dosage according to the classification snakebite envenomation, the rural or urban origin of the patient, prehospital nonmedical management received, and the age of the patient were defined as potential confounding variables.

Results are expressed as crude and adjusted relative risks for each of the outcome variables. Results were assumed to be statistically significant if the *P* value was <0.05.

The statistical software R (version 4.1.0, R Foundation for Statistical Computing, Vienna, Austria) and R Studio (version 1.4.1717, R Studio Inc., Boston, United States) were used to analyze the data [18].

## Results

364 clinical records of patients admitted to both hospitals were reviewed. A total of 50 patients were excluded, mainly due to the inability to determine the time between the snakebite and antivenom administration (Fig 1). Finally, 304 patients with a snakebite by *Bothrops spp.* was analyzed, 54.6% (*n=*166) in the antivenom administration early group and 45.4% (*n=*138) in the late group.

**Fig 1.**
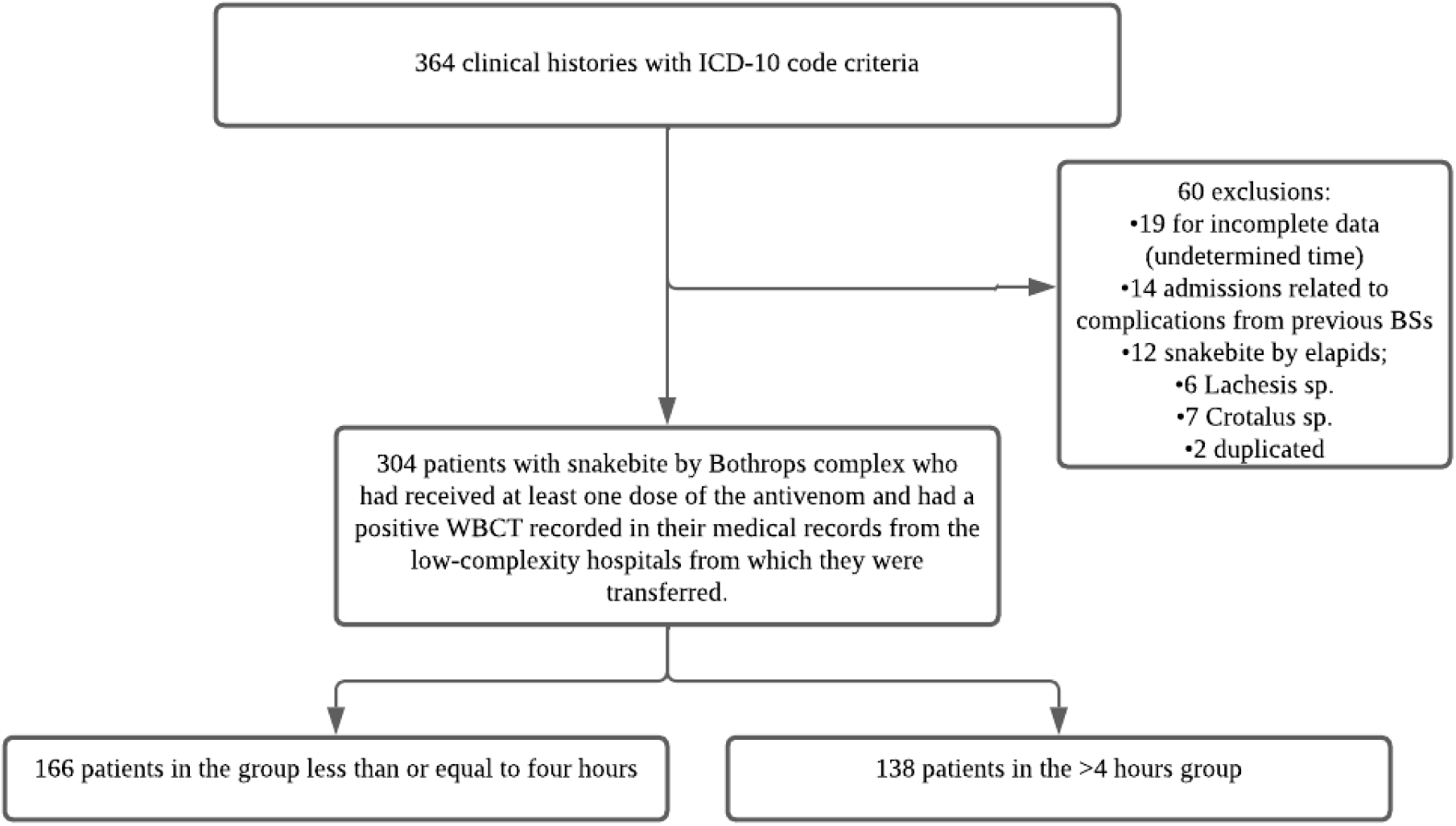
Recruitment flowchart of cohort participants. Figure created by the authors.

The median age was 35 years (IQR 18-53); 79.0% (n=241) of patients were men, were 89.8% (*n=*274) from rural areas; and 53.8% (*n=*164) were farmers. A total of 86.5% (n=276) of the patients belonged to the subsidized health regime. Table 1 describes the socio-demographic, clinical variables of the population.

**Table 1.** Sociodemographic, clinical and care characteristics.

| Variable | Group ≤ four hours (n=166) | Group > four hours (n=138) | Total (n=304) |
| --- | --- | --- | --- |
| Age (years), median (IQR) | 36 (19-53) | 35 (17-51) | 35 (18-53) |
| Profession, n (%) |  |  |  |
| Farmer | 96 (57.8) | 68(49.3) | 164 (53.8) |
| Male sex, n (%) | 141 (84.9) | 100 (72.5) | 241 (79.0) |
| Rural residence, n (%) | 144 (86.7) | 129 (93.5) | 274 (89.8) |
| **Background, n (%)** |  |  |  |
| Diabetes | 3 (1.8) | 2 (1.4) | 5 (1.6) |
| Kidney disease | 4 (2.4) | 1 (0.7) | 5 (1.6) |
| Hypertension | 11 (6.6) | 3 (2.3) | 14 (4.5) |
| **Envenomation description** |  |  |  |
| Local envenomation |  |  |  |
| Mild | 49 (29.5) | 37 (26.8) | 87 (28.5) |
| Moderate | 63 (38.0) | 55 (39.9) | 118 (38.7) |
| Serious | 54 (32.5) | 46 (33.3) | 100 (32.8) |
| Systemic envenomation |  |  |  |
| Mild | 96 (57.8) | 44 (31.9) | 140 (45.9) |
| Moderate | 33 (19.9) | 46 (33.3) | 79 (25.9) |
| Serious | 37 (22.3) | 48 (34.8) | 86 (28.2) |
| Number of ampules applied in LH, (median, min-max) | 4 (1-12) | 6 (2-12) | 6 (1-12) |
| Number of ampules applied in HCH, (median, min-max) | 4 (1-12) | 4 (1-28) | 4 (1-28) |
| Additional doses, n (%) | 78 (47.0) | 56 (40.6) | 73 (24.2) |
| Insufficient dosage, n (%) | 11 (6.6) | 8 (5.7) | 19 (6.2) |
| Time in minutes for antivenom application after the SB, median (IQR) | 120 (90-180) | 990 (420-1702) | 240 (120-845) |
| **Clinical manifestations upon admission to the HCH** |  |  |  |
| Heart Rate (beats per minute) | 82 (73-95) | 88 (76-99) | 85 (75-98) |
| Mean arterial pressure (mmHg) | 90 (83-97) | 90 (81-98) | 90 (82-98) |
| Hemorrhagic or clear blisters, n (%) | 26 (15.6) | 34 (24.6) | 60 (19.6) |
| **Laboratory values in the first 24 hours (IQR)** |  |  |  |
| Hemoglobin (g/dl) | 14.4 (13.0-15.7) | 13.8 (12.2-15.4) | 14.2 (12.6-15.5) |
| Creatinine (mg/dl) | 0.9 (0.7-1.0) | 0.9 (0.6-1.1) | 0.9 (0.7-1.1) |
| Platelets (10<sup>3</sup> cel/mcl) | 238 (184-277) | 213 (174-282) | 227 (179-278) |
| TP (seconds) | 15.0 (11.5-14.5) | 15.1 (13.0-24.0) | 15.1 (12.9-19.5) |
| INR | 1.3 (1.1-1.8) | 1.3 (1.1-2.1) | 1.3 (1.1-1.7) |
| CK (mcg/l) | 186 (128-414) | 188 (120-370) | 187 (120-383) |
| **Laboratory values, 25-48 hours after the snakebite (IQR)** |  |  |  |
| Hemoglobin (g/dl) | 13.0 (11.8-14.5) | 12.3 (10.1-13-7) | 12.7 (11.3-14.2) |
| Creatinine (mg/dl) | 0.8 (0.6-1.0) | 0.9 (0.6-1.0) | 0.8 (0.6-1.0) |
| Platelets (10<sup>3</sup> cel/mcl) | 188 (158-246) | 172 (141-223) | 180 (150-235) |
| TP (seconds) | 12.9 (11.6-14.5) | 13.2 (12.3-16.2) | 13.1 (11.7-14.8) |
| INR | 1.1 (1.0-1.3) | 1.2 (1.1-1.4) | 1.2 (1.0-1.3) |
| CK (mcg/l) | 275 (88-550) | 175 (98-482) | 231 (90-491) |
| **Laboratory values, more than 48 hours after the snakebite (IQR)** |  |  |  |
| Hemoglobin (g/dl) | 12.7 (10.7-14.0) | 11.7 (9.9-13.3) | 1.7 (9.0-13.3) |
| Creatinine (mg/dl) | 0.8 (0.6-0.9) | 0.8 (0.6-1.0) | 0.8 (0.6-1.0) |
| Platelets (10<sup>3</sup> cel/mcl) | 178 (150-246) | 188 (118-265) | 180 (131-264) |
| TP (seconds) | 12.1 (11.3-13.2) | 12.4 (11.8-13.8) | 12.3 (11.5-13.4) |
| INR | 1.0 (1.0-1.1) | 1.1 (1.0-1.2) | 1.1 (1.0-1-2) |
| CK (mcg/l) | 409 (85-1229) | 137 (87-430) | 212 (86-714) |

Continuous variables are expressed as medians and Interquartil Range, unless otherwise indicated. IQR: Interquartile range; SB: Snakebite; LH: Local hospital; HCH: High-complexity hospital; HR: heart rate; MAP: Mean arterial pressure in millimeters of mercury; PT: Prothrombin time; mcg/lt: micrograms per liter; g/dl: grams per deciliter; mg/dl: milligrams per deciliter; cell/mcl: cells per microliter; CK: Total creatine kinase; INR International Normalized Ratio

Antivenoms used for the treatment of the snakebite envenomation were Probiol® (3.2% [*n=*10]), Bioclon® (13.4% [*n=*41]), National Health Institute of Colombia (53.9% [*n=*164]), and not registered in 28.9% (*n=*88) of the cases. The time interval between the snakebite and the administration of the antivenom behaved non-normally in both groups (p <0.05).

Persistence of VICC between 25–72 hours was different between groups, in the late 10.8% (*n=*24) meanwhile in the early 5.4% (*n=*14) (*P* value =0.02). In patients with persistence of VICC, the median level of INR was 1.70 (1.59-2.06), in the early group 1.70 (1.55-2.06) and in the late 1.68 (1.54-2.10). Behavior of INR in these patients is illustrated in Fig 2.

**Fig 2.**
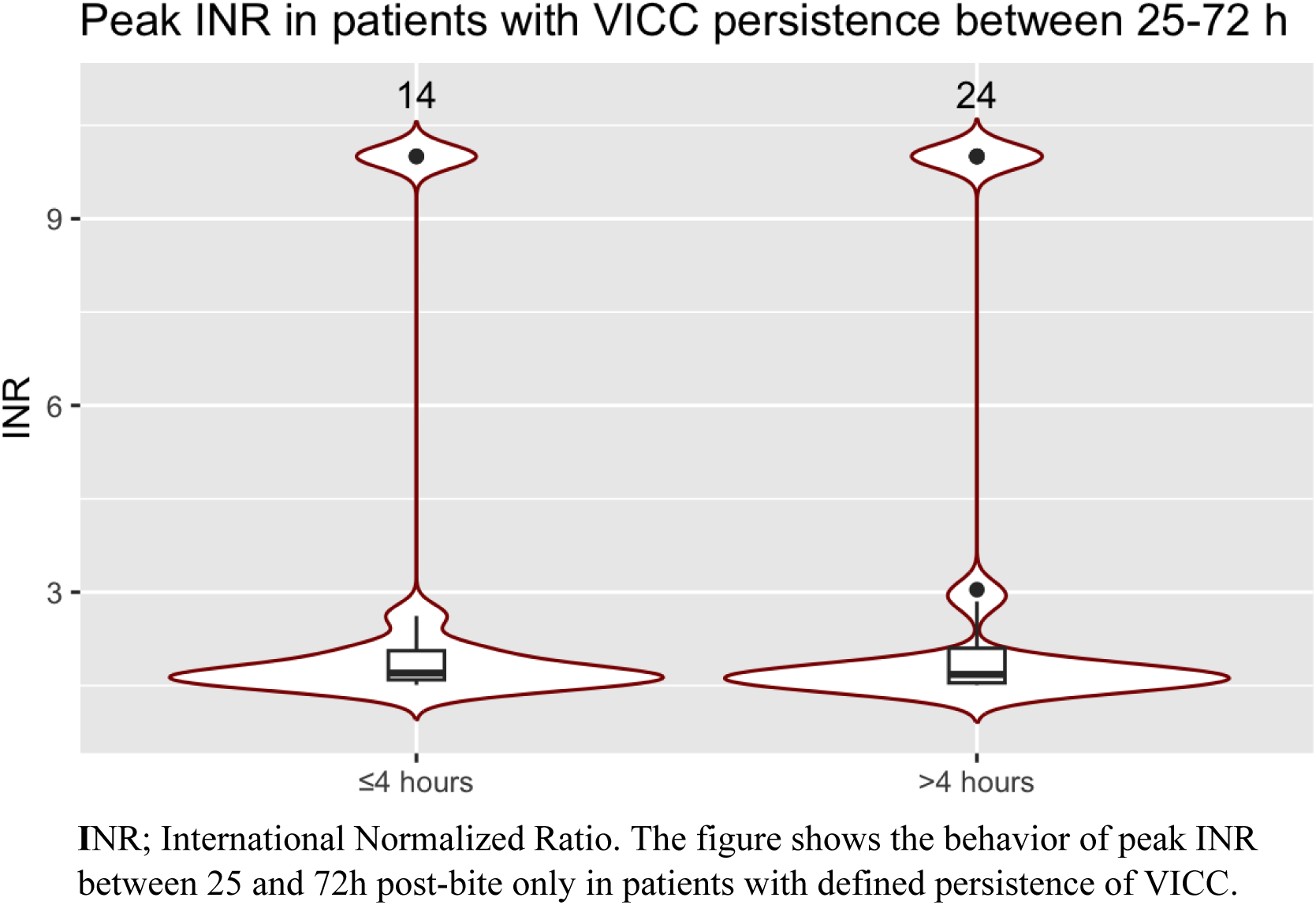
International Normalized Ratio (INR) by time points in defined groups.

Clinical bleeding occurred in 24.3% (*n=*74) of the population, being more frequent in the late compared with the early group (33.3% [*n=*46] and 16.8% [*n=*28], respectively), as well as major bleeding (early group 3.6% [*n=*6] *vs.* late 10.8% [*n=*15], P value =0.02) (Table 2). Digestive bleeding was the most frequent hemorrhagic manifestation in the 10.1% (*n=*31) of the entire population (Table 3).

**Table 2.** Absolute and relative frequencies of clinical outcomes in patients with a snakebite envenomation.

| Variable | Group $\leq 4$<br>hours ($n=166$) | Group $> 4$<br>hours ($n=138$) | Total ($n=304$) |
| --- | --- | --- | --- |
| **Clinical Outcomes, n (%)** |  |  |  |
| **Persistence of VICC between 25 and 72 hours** | 14 (8.4) | 24 (17.3) | 38 (7.8) |
| **Clinical Bleeding** | 29 (17.0%) | 47 (31.8%) | 76 (23.8%) |
| **Rhabdomyolysis** | 17 (9.9%) | 18 (12.1%) | 35 (10.9%) |
| **Acute kidney injury** | 27 (15.9%) | 33 (22.9%) | 60 (18.8%) |
| **Days spent in SCU/ICU; median (IQR)** | 0 (0) | 0 (2.1) | 0 (1) |
| **Death** | 3 (1.8%) | 2 (1.4%) | 5 (1.5%) |

VICC: Venom-induced consumption coagulopathy; SCU/ICU: Special/Intensive Care Unit.

**Table 3.** Absolute and relative frequencies of Hemorrhagic manifestations in patients with a snakebite envenomation.

| Variable | Group ≤ 4<br>hours (n=166) | Group > 4<br>hours (n=138) | Total (n=304) |
| --- | --- | --- | --- |
| **Hemorrhagic manifestations, n (%)** |  |  |  |
| **Epistaxis** | 1 (0.6%) | 4 (2.8%) | 5 (1.6%) |
| **Digestive bleeding** | 12 (7.2%) | 20 (14.4 %) | 32 (10.5%) |
| **Hematuria** | 8 (4.8%) | 14 (10.1%) | 19 (7.2%) |
| **Gingivorrhagia** | 5 (3.0%) | 2 (1.4%) | 7 (2.3%) |
| **Central nervous system** | 2 (1.2%) | 4 (2.8%) | 6 (1.9%) |
| **Bite site** | 0 (0%) | 2 (1.4%) | 2 (0.6%) |
| **Venipuncture site** | 0 (0%) | 1 (0.7%) | 1 (0.3%) |

Relative risk for persistence of VICC was greater in the late group (2.06 [95% IC 1.11-3.83]), as well as for clinical bleeding and major bleeding (1.97[1.30-2.98]; 3.00 [1.19-7.54], respectively) (Table 4). The length of stay in the special/intensive care unit, rhabdomyolysis and death rate was no different in both groups. Estimated relative risks with its Confidence Intervals for other explored outcomes are presented in Table 4.

**Table 4.**
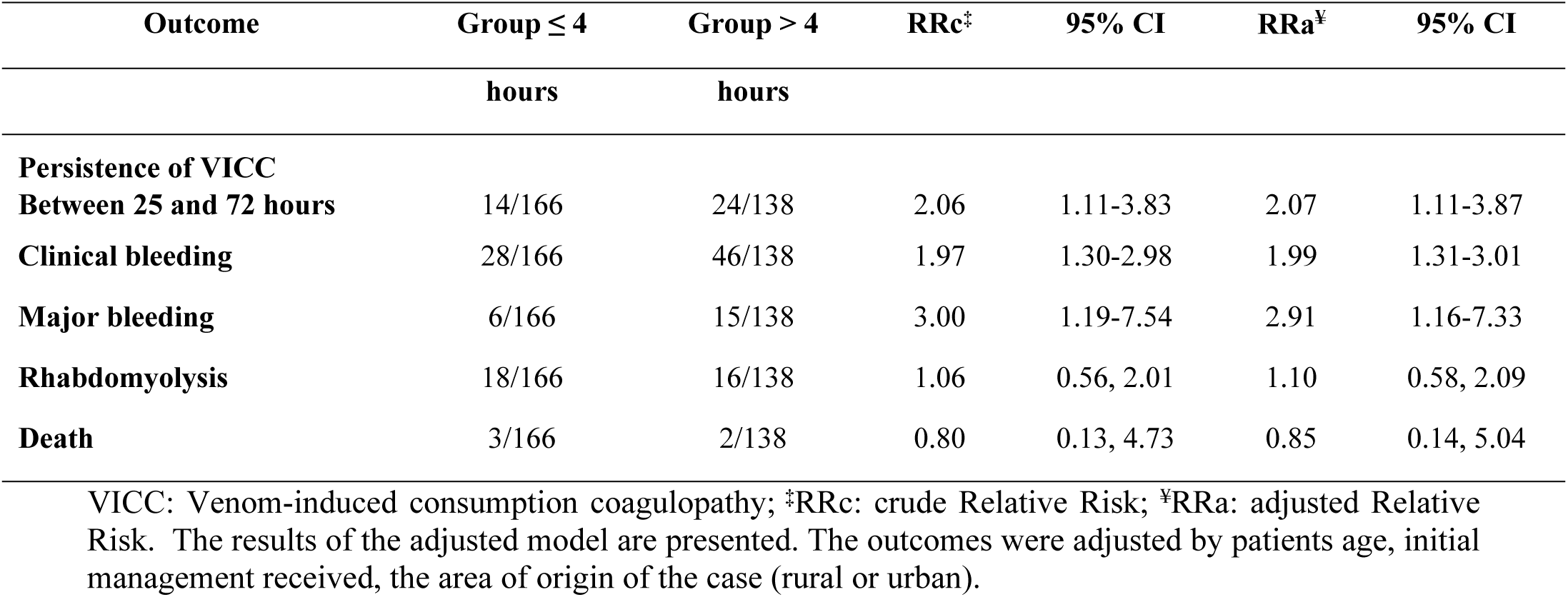
Relationship between the time of application of antivenom and the development of clinical important outcomes.

## Discussion

In the current study, we evaluated the association of the late administration of antivenom serum and the presentation of VICC and other related outcomes in patients with snakebite envenomation by *Bothrops spp*. Our results show a significantly persistence of VICC, clinical bleeding and major bleeding in patients who received the antivenom serum more than five hours after the snakebite, compared with those who received it early.

Results of studies in relation to a time-dependent phenomenon of the antivenom administration are varied. In a prospective observational study; Olivera Pardal *et al.* [19] compared in an open clinical trial in Brazil the effectiveness of antivenom for the *Bothrops-Lachesis* genera versus an specific one for the species *B. atrox - L. muta*, for the resolution of coagulopathy, they found that 86% of patients reversed coagulopathy and clinical bleeding stopped in almost all patients six hours after antivenom administration. They used the 20-min WBCT or the clinical resolution of bleeding, as the definition of coagulopathy. Time intervals from the snakebite to the antivenom administration were longer. Snake species analyzed in this trial share similarity with the species involved in the current study.

Also, in an observational study, Silva et al. compared patients who developed VICC after a snakebite by *D. ruselli*, and receive antivenom and those who did not[8]. The results showed that the group that did not receive antivenom had lower VICC recovery, but 21% of patients in the antivenom group still with prolonged INR at 48 hours. In the same way, Mion et al., found that patients with snakebite envenoming from *Echis spp*. that received antivenom serum had a more rapid recovery of PT, but a proportion of patients near 20%, persisted with prolonged PT between 48 and 72 hours after treatment with antivenom [6]. Therefore, not all patients who developed VICC following a snakebite exhibit resolution of the same within 48 hours.

Finally, Houcke *et al.*, in a cohort study evaluating the effect of early and late administration of antivenom (six-hour time point definition) to patients with snakebite envenomation, including *Bothrops spp.*, found a slower recovery of INRs in the late group[9].

In contrast, Johnston et al.[20], observed that VICC resolution was not associated timely antivenom administration, however they did observe a lower frequency of neurotoxicity, myotoxicity, acute kidney injury and intubation in patients with *Oxyuranus spp*. snakebite when antivenom was administered within four hours of the bite. In the same way, Isbister et al. [21], found no differences in recovery of severe VICC among patients who suffered a snakebite by Australian elapid and received antivenom ≤6 hours, compared with those beyond >6 hours.

Our findings can be explained given the inter and intra-species variability in venom composition compared to those in literature. Not all snakebites have the same behavior in relation to the development of hemotoxicity and coagulopathy[7], which also, is related to factors such as geographic location, but also the definition of VICC used, such as described above studies, that is reflected in the diverse incidence of VICC [2], [5], [16], [22], [23].

In Colombia, low-complexity hospitals generally lack access to PT/INR, and most of patients receives antivenom in conjunction with WBCT results prior to conducting coagulation tests [23], [24]. This practice is based in the recommendations outlined in the WHO and the Ministry of Health of Colombia [12], and can explain the low frequency of VICC of our study.

On the other hand, our data show almost twofold the increased risk of developing clinical bleeding and almost threefold increased risk of developing major bleeding in the late group. VICC is a clinical important and prevalent complication arising from snakebite envenomation. It can induce severe hemorrhagic manifestations within the gastrointestinal tract or the central nervous system, with the potential to be life-threatening, and therefor the importance of performing interventions to reduce the clinical risk of bleeding [6], [16].

These findings are similar to those presented in study of Otero et al. [10] conducted in 2002. who describes patients with a snakebite by the genus *Bothrops, Porthidium* and *Bothriechis*. The analysis revealed an increased frequency of bleeding in genitourinary and gastrointestinal systems. Furthermore, the study showed that patients who received antivenom more than 2 hours after the snakebite had a higher incidence of central nervous system bleeding, like our study.

Mortality risk was not different between groups, which could be explained by the low frequency of the event, resulting underrepresented. Scientific and non-governmental reports have shown how the risk of death from all causes increases when antivenom is not administered in a timely manner [1], [3]. For this reason, is crucial the urgent need for accessibility to antivenoms [3], [25].

Non-medical prehospital management in addition to the greater frequency of the event in rural areas, distant to health facilities, could have contributed to the delay in receiving the antivenom. The results of the cohort of Silva et al.[8], showed that one of the main factors of delay for consultation after the snakebite was the absence of symptoms in 50% of the cases, and a preference for non-medical treatments in 14% of the cases [8].

The findings in the present study could contribute to the development of strategies for improving the timeliness of care, specifically in the administration of antivenom to patients with a snakebite in the Colombian context [26].

Given the retrospective nature of our research, data related to the patient’s memory of snakebites, and the quality of the data from the referring hospital, may have been imprecise, however, for primary and secondary outcomes, the variables were all objective and abstracted without clinical interpretation or inference. Also, some variation might be expected in the time intervals between laboratory tests after hospital admission, however, such variation should not be expected to be different between both groups.

## Conclusion

The late antivenom administration increased risk of persistence of VICC, clinical and major bleeding in patients with snakebite envenomation by Colombian *Bothrops spp*.

## Data availability statement

Data is available for public consultation, as Supporting Information, as S1 Appendix.

